# Refined Matrix Completion for Spectrum Estimation of Heart Rate Variability

**DOI:** 10.1101/2022.10.30.22281728

**Authors:** Lei Lu, Tingting Zhu, Ying Tan, Jiandong Zhou, Lei Clifton, Yuan-Ting Zhang, David A. Clifton

## Abstract

Heart rate variability (HRV) is the reflection of physiological effects modulating heart rhythm. In particular, spectral HRV metrics provide valuable information to investigate activities of the cardiac autonomic nervous system. However, uncertainties and artifacts from measurements can reduce signal quality and therefore affect the evaluation of HRV measures. In this paper, we propose a new method for HRV spectrum estimation with measurement uncertainties using matrix completion (MC). We show that missing values of HRV spectrum can be efficiently estimated using the MC method by leveraging the low rank property of the spectrum matrix. In addition, we proposed a refined matrix completion (RMC) method to improve the estimation accuracy and computational efficiency by introducing model information for the HRV spectrum. Experimental studies on five public benchmark datasets show the effectiveness and robustness of the developed RMC method for estimating missing entries for HRV spectrum with different masking ratios. Furthermore, our developed RMC method is compared with five deep learning models and the traditional MC method; the results of this comparison study demonstrate that our developed RMC method obtains the least estimation error with the minimal computation cost, indicating the advantages of our developed method for HRV spectrum estimation.

## I. Introduction

Heart rate variability (HRV) is defined as the changes of time intervals between consecutive heartbeats [1]. Modulated by the sympathetic and parasympathetic branches of the autonomic nervous system (ANS), these changes are considered as reliable reflections of many physiological factors modulating the normal rhythm of the heart [2], [3]. For example, high HRV suggests healthy cardiac activities, as it allows a better adjustment to external and internal stimuli; In contrast, low HRV indicates inadequate parasympathetic or excessive sympathetic activity, which is believed to be associated with increased morbidity [4], [5]. HRV is therefore widely used to assess physiological and pathological conditions, particularly in the setting of cardiovascular disease (CVD) [6], [7], which is currently the most common cause of death worldwide, with an estimation of 30% of global mortality according to the World Health Organization (WHO) [8], [9].

HRV data can be evaluated using a variety of signal analysis methods. For example, HRV measures can be calculated in the time domain, which typically computes statistical indices of HRV data, including the mean value of normal-to-normal RR intervals (NN), root mean square of successive difference of intervals (RMSSD), and standard deviation of NN intervals (SDNN) [10], [11]. Compared to time domain analysis, HRV indices in the frequency domain describe absolute or relative power distribution with different frequency bands of the HRV data. In particular, spectral features of HRV data were found to be reliable markers of sympathetic and/or parasympathetic activity [12]. For example, the low frequency (LF: 0.04 - 0.15 Hz) of HRV spectrum is generally mediated by sympathetic and parasympathetic actions; while the high frequency (HF: 0.15 - 0.4 Hz) is mediated by the parasympathetic nervous system; as a consequence, the LF/HF ratio can be used as an index of sympathovagal balance [13], [14].

The most common method used to derive HRV data is via electrocardiography (ECG) recordings. With appropriate QRS detectors, R-peaks in the ECG morphology can be identified, then HRV sequence can be obtained by computing the RR intervals [15]. Other than using ECGs, some studies suggest using photoplethysmography (PPG) to derive the HRV data, where PPG is a non-invasive technique that utilises optical principles to obtain pulse waves from the microcirculation in peripheral tissue [15], [16]. In particular, the use of PPG signal to detect pulse-wave related HRV, also known as pulse rate variability (PRV), has attracted considerable attentions in recent years [16], which is due to the convenience and low cost of acquiring PPG signals using wearable devices [17].

Despite the widespread use of ECG or PPG signals for HRV analysis, it is still challenging to obtain reliable HRV data from these signals. This can be due to a number of factors; (*i*) either the ECG or PPG signal is sensitive to measurement noise and uncertainties, e.g., missing values, which would have subsequent effects on the HRV data [17]; (*ii*) the analytical approach and computational procedures present complexities when deriving HRV measures using these signals, such as R peak identification using QRS detectors, RR interval interpolation and resampling to generate the tachogram. These processes could introduce additional estimation errors on HRV data analysis [15]; and (*iii*) the ECG signal is a composition of multiple frequency components that are produced by a variety of physiological processes, and abnormalities in these processes can easily affect the frequency spectrum of HRV data [10].

To this end, many computational techniques have been developed to address these challenges in the analysis of HRV data. For example, the Gaussian model was used to estimate frequency components from noisy HRV data [18]; ensemble machine learning was used to explore linear and non-linear correlations between HRV features derived from ECG data [19]; and a hybrid deep learning model was developed to combine different HRV features and produced reliable classification performance [20]. Nevertheless, machine learning methods, and in particular deep neural networks, typically require expertise in model development and hyperparameters tuning. Furthermore, it is essential to form a large number of training samples to guarantee the model performance. Therefore, more efficient and reliable methods need to be developed to address these uncertainties in HRV data analysis.

Low-rank matrix completion (MC) is a promising method to estimate missing entries for incomplete or inexact observed data [21], where the low-rank property of data measurements is believed to exist in many real-world applications. This is due to the characteristics of intrinsic low dimensional space or an underlying trend of massive measured data [22], [23]. Using the low-rank property, missing entries in the observations can be potentially inferred by the partially sampled data. The MC technique has been studied for image processing [22], remote sensing [24], and wireless sensor networks [25]. It also has been used in bioinformatics, such as in the recognition of long non-coding RNA (lncRNA) disease associations [26], and in microRNA target prediction [27]. However, the use of MC method for HRV spectrum estimation is largely unexplored in the literature.

In this paper, we develop a new method to estimate uncertainties of HRV spectrum using the MC method. By leveraging the low-rank property of power spectral density (PSD) of HRV data, we show that uncertainties in the spectrum can be efficiently inferred using partial entries of the data matrix. In particular, we investigate characteristics of the HRV data; i.e. LF and HF of the spectrum, and a new framework of refined matrix completion (RMC) method is developed for missing value estimation by using important data entries in the HRV matrix, which is expected to have more efficient and robust performance on the uncertainty estimation. To evaluate the performance of our method for HRV spectrum estimation, extensive experimental studies are implemented on five benchmark datasets; In addition, the traditional MC method and five deep recurrent models are used for comparison study with our developed RMC method. The experimental results and comparison studies demonstrate the robustness and advantages of our developed method for HRV spectrum estimation. The contributions of this paper are:

1. A new method for uncertainty estimation of HRV spectrum is developed by using the MC technique, which leverages the low-rank property of data matrix that is derived from HRV spectrum.
2. To improve the estimation performance and computation efficiency, a new framework with refined matrix completion is proposed by investigating the modelled spectrum of HRV data.
3. Extensive experimental studies are implemented on five well-known ECG benchmark datasets to investigate the performance of our developed model for HRV spectrum estimation.
4. Comparative studies with five different types of deep recurrent neural networks and the traditional MC method are performed to show advantages of our developed model for missing entry estimation.

The remainder of this paper is organised as follows. Section II formulates the problem of HRV spectrum estimation. Section III develops the MC method and refinement. Section IV performs the analysis of HRV data. The results of spectrum estimation and comparison study are presented in Section V, and finally conclusions are drawn in Section VI.

## II. Problem Formulation

### A. Notations

In this paper, the set of real numbers is denoted as ℝ, and the set of natural numbers is denoted as 𝒩. Bold uppercase letter **A** = {*a*_*i,j*_}_*i*=1,…,*n,j*=1,…,*p*_ *∈ ℝ*^*n×p*^ denotes a matrix with *n* rows and *p* columns. The notion 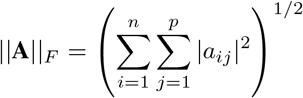 denotes its Frobenius norm. The notion ||**A**|| _*_ = Σ _*i*_ *σ*_*i*_(**A**) is the nuclear norm of this matrix where *σ*_*i*_(**A**) denotes the *i*^*th*^ singular value of **A**, and its 2-norm is defined as ||**A**|| _2_ = *σ*_*max*_(**A**) where *σ*_*max*_ is the largest singular value.

### B. Problem Formulation

Assuming the sequence *z*(*t*) *∈* ℝ, *t ∈* [0, *Ts*] is the pre-processed HRV data with its frequency components represented as

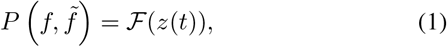

where 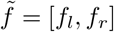 indicates the interested frequency range of the HRV data, *f*_*l*_ and *f*_*r*_ are the left and right bounds separately. Here ℱ (·) denotes the spectrum operator projected to the interested frequency range 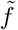, and 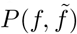 is the calculated power spectral density (PSD) function of the HRV data. From the PSD signal 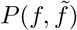, we can extract different frequency variables that are related to the PSD. More precisely,

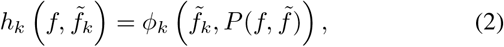

where *k* = 1, 2, · · ·, *N*_*k*_ indicates different types of HRV variables, such as the power of LF or HF, or the ratio between the LF and HF. The nonlinear mapping *ϕ*_*k*_(·,·) computes frequency variable *h*_*k*_ from the spectrum *P* (·,·) for the *k*^*th*^ type of HRV variables in the 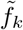 frequency range.

We note that the measurements of HRV data depend on the quality of ECG recordings, which may be prone to measurement noise and missing values. Other factors such as the respiration rate, circadian rhythms, and physiological states may also have impact on quality of ECG signals [10]. These uncertainties will then have subsequent influence on the frequency analysis of HRV data, and therefore it is challenging to accurately estimate the HRV spectrum.

Assuming the data from *N*_*s*_ subjects have been collected, and a matrix 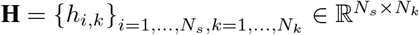 can be derived from the measurements, which is defined as

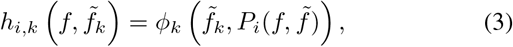

where *P*_*i*_ is the PSD of the *i*^*th*^ subject. It is noted that the PSD of HRV data usually identifies dominating frequency components with amplitudes of many non-dominating frequencies that are small enough to be negligible, leading to a sparse matrix; In addition, frequency components of the PSD spectrum can be potentially affected by measurement noise, and computational procedures of HRV data analysis may also produce uncertainties in the spectrum. In this work, the PSD data derived from large amount of collected ECGs demonstrate similarities among the spectrum waveforms, indicating the low rank property of the spectrum matrix. This motivates us to use low-rank matrix completion to perform the matrix approximation for uncertainty estimation. In particular, we propose the refined matrix completion (RMC) to estimate uncertainties in a more efficient way.

## III. Methodology

This section introduces the concept of low-rank matrix completion for a sparse matrix. The key idea is to find a good approximation of the matrix, with respect to some pre-defined cost function such as finding an optimal approximation by minimizing the rank of the matrix. The section starts from a revisit of low-rank matrix completion, followed by the matrix approximation with interested zone, and then the introduction of the refined matrix completion.

### A. Low-rank Matrix Completion

Assuming matrix 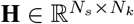 is sparse, i.e., only a limited number of entries in the matrix are non-zero. We denote the set Ω as the collection of non-zero entries of the matrix **H**, which is represented as

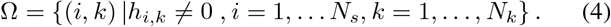

It is noted that the spectra **H** derived from data segments usually are obtained by recursively slicing the HRV sequence, which indicates the existence of redundant information in the matrix. In order to keep useful information and reduce the redundant information, a low-rank matrix completion technique [21], [28] is adapted in this paper. It can be formulated by approximating matrix **H** in terms of the smallest rank, i.e.,

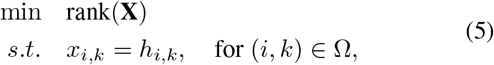

where matrix 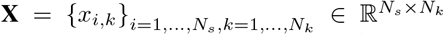an optimal approximation matrix that can keep the majority information of **H**.

The optimisation of rank-minimization problem in Eq. (5) is generally computationally intractable. Instead of solving the original problem of rank norm, a convex relaxation based on the nuclear norm is given by [21],

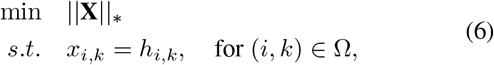

where, || · ||_*_ indicates the nuclear norm of matrix **X**, which is defined as the summation of its singular values. In particular, it is demonstrated that a smaller nuclear norm indicates the lower rank of the matrix [21]. However, it is generally too harsh to force the estimation of all observed entries in the matrix, so alternatively, a more practical approximation of Eq. (6) can be formulated by relaxing the constraint [29], [30].

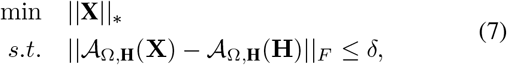

where || · ||_*F*_ denotes the Frobenius norm, *δ* is a small value to constrain the estimation, and 𝒜_Ω,**H**_(·) is a projection operator that is defined as

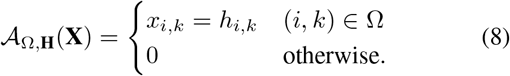

The optimisation problem in Eq. (7) can be formulated as a penalised least-squares convex program [31], which can be efficiently solved by the singular value thresholding (SVT) technique as described in [32].

### B. Matrix Approximation with Interested Zone

The above-mentioned method assumes that observed entries including the missing ones should be random variables satisfying the uniform distribution [21]. However, we note that some frequency components of HRV spectrum are prone to measurement certainties. For example, the LF spectrum could be affected by respiration rate, and HF spectrum would be affected by measurement noises. In this study, we investigate using some frequency components of the spectrum to estimate noise affected entries. This indicates that some parts of columns or rows (instead of random entries) are missing. Therefore the traditional low-rank MC techniques mentioned above cannot be applied directly. Alternatively, a method called Interest-Zone Matrix-Approximation (IZMA) [28] is proposed to solve the low-rank MC problem when some columns/sub-columns or rows/sub-rows are missing. More specifically, the IZMA tries to solve the following optimization problem,

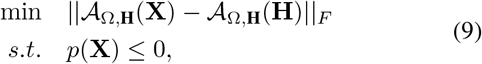

where the nonlinear mapping 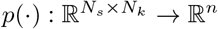 is a smooth function for some *n* ∈ 𝒩. For example, *p*(**X**) can be some constraints with respect to the estimated matrix, such as *p*(**X**) = ||**X**||_2_ *− λ, λ >* 0. Another example of this nonlinear mapping is *p*(**X**) = ||**X**|| _*_ *−λ* for some *λ >* 0 as described in [28].

The optimisation problem with entries in an interested zone in Eq. (9) can be solved by the following steps; It first approximates the following optimisation for the constrained full matrix,

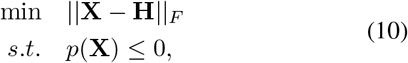

The solution to the optimisation in Eq. (10) with the spectral norm constraint can be estimated by,

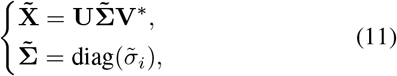

where matrices **U** and **V** are calculated by **H** = **UΣV**^*^, with **Σ** = diag(*σ*_1_, *σ*_2_, · · ·, *σ*_*n*_). Next, the diagonal matrix is thresholded as 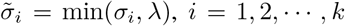,*i* = 1, 2,, *k*, and the approximation 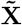 can be obtained by the reconstruction.

Then, the estimation of matrix **H** with entries in the interested zone can be updated as follows,

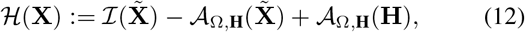

where ℐ is an identity operator, and ℋ (·) denotes the updating process, which combines the known entries in matrix **H** and the estimated entries in matrix 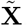.

Next, the estimated matrix is replaced with ℋ (**X**) and the iteration is repeated starting from Eq. (10). The estimation is evaluated by *ϵ*(**X**) ≜||𝒜 _Ω,**H**_(**X**) *−*𝒜_Ω,**H**_(**H**)||_*F*_, and missing entries in matrix **H** can be approximated until *ϵ*(**X**) converges. For convenience, we denote the whole iteration process of the constrained approximation as 𝒯 (**H**, *λ*), which will be used in the next section for the HRV spectrum estimation.

It is noted that the matrix approximation in Eq. (9) differs from the low-rank MC as presented in Eq. (5), where the low-rank MC requires to fix the known entries for the estimation, while the matrix approximation in Eq. (9) minimize the objective function with Frobenius norm to the constraint function. In particular, the technique of matrix approximation with an interested zone can also be extended as matrix completion, which develops a new iteration process separate from the traditional SVT technique; more details of the iteration process can be found in [28], and it will also be discussed in Section III-C.

The above approximation of missing entries is data-driven and implemented using an iteration process. We note that for the analysis of HRV data, some mathematical techniques can be used to model the spectrum, such as the Gaussian model as pointed out in [18], the model can be used to characterise HRV spectra and investigate the relationship between them. In the next subsection, we will use the Gaussian model to develop a refined matrix completion (RMC) by generating a new matrix with a much lower dimension. Consequently, this new matrix is employed to perform low-rank matrix completion by using techniques such as IZMA, which is expected to improve the efficiency and performance of the estimation.

### C. Model-based Refined MC for HRV Spectrum Estimation

In general, the spectrum of HRV data can be divided into different frequency bands with specific characteristics, such as the Mayer wave in the LF spectrum, and the RAS wave in the HF spectrum. It is assumed that the matrix 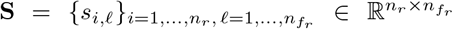 consists of *n*_*r*_ spectra that are derived from the HRV data, where 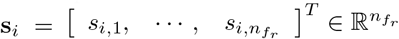 is a vector of the spectrum. Given the relationship between the LF and HF components of the spectrum [18], we assume the LF part can be used as a reference to estimate uncertainties in the HF part, or the HF can be used to estimate the LF part. Specifically, for each **s**_*i*_ *∈* **S**, it can be modelled with Gaussian functions [18], and the entry *s*_*i,ℓ*_ can be represented as follows,

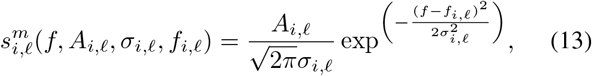

where *f ∈* [*f*_*l,i*_, *f*_*r,i*_] indicates the interval of frequency band of interests, i.e., LF or HF spectrum. Consequently, *A*_*i,ℓ*_ *>* 0 is the weight of amplitude, *f*_*i,ℓ*_ *>* 0 is the peak position of frequencies in the spectrum, and *σ*_*i,ℓ*_ *>* 0 is its standard deviation for this frequency. For simplicity of notations, 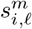 is used to represent 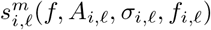 when no confusion arises, and 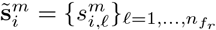 indicates the modelled spectrum.

It is noted that the HRV spectrum can be characterised with different frequency components, and they can be simulated by changing the frequency intervals of the modelling, after which the complete spectrum can be obtained by combining these different frequency components. Next, we use the modelled information to define a new matrix for the estimation of missing entries. For the convenience of presentation, it is assumed that some entries of matrix **S** are missing, and the set 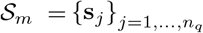 contains the rows in **S** with missing values.

For any **s**_*j*_ *∈ S*_*m*_, we model the spectrum and obtain 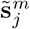 with Eq. (13), and then calculate the correlation between 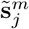 and 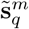 spectra, where 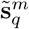 is the modelled information of spectrum in **S**, and the correlation is computed as follows [33],

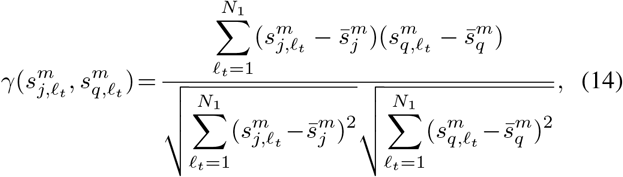

where 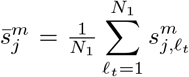 and 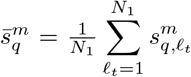 are means of the two spectra respectively, 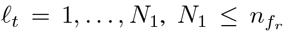 is the index of known components of the spectrum, and 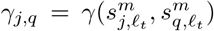 indicates the relationship between the two spectra.

After calculating *γ*_*j,q*_ for the **s**_*j*_ spectrum, a new vector 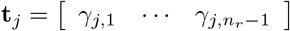 can be obtained, we sort **t**_*j*_ and select the *K* highest ranked elements for **s**_*j*_, then a set that contains the identified indices can be formed as,

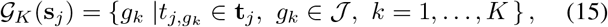

where 𝒥 = 1, …, *n*_*r*_ *−*1 denotes the index of spectrum in matrix **S**.

Next, a refined matrix 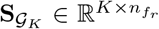 can be formulated by combining **s**_*j*_ and the identified HRV spectra. Consequently, the optimisation in Eq. (9) can be updated as,

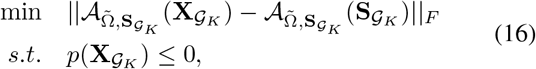

where 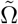 is the set of observed entries in the refined matrix, 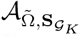 is the updated operator to model the missing values, 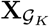 indicates the estimated matrix, and the spectral norm 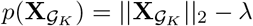can be used for the constraint.

For the estimation of missing entries in the refined matrix, we first approximate the following constrained full matrix,

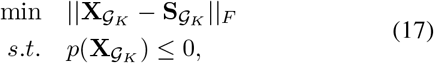

where the soft imputing method as described in Eq. (11) can be used to solve the estimation in Eq. (17). Then, by updating the parameter *λ* in the constraint function, missing entries in the refined matrix can be estimated using the iteration process as described in Section III-B. Algorithm 1 presents the pseudo code of the proposed RMC method. The hyperparameters tolerance *e*_*tol*_ and *λ*_*tol*_ are set as 10^−8^ and 10 according to the configuration in [28].

As an illustration, Figure 1 demonstrates the concept of estimating missing entries for HRV spectrum using the RMC technique, where the matrix is constructed with *m* spectra that are derived from HRV data segments. The *x*-axis of the matrix indicates the frequency band of interests, and *y*-axis shows the indices of data segments. Suppose the data of **s**_3_ spectrum are affected by measurement uncertainties, which are masked as missing entries. We model the spectra in the matrix, and use the LF spectrum as references to identify relevant elements. Then, these identified spectra, such as vectors **s**_2_, **s**_5_, and **s**_*m*_, along with **s**_3_ form a new matrix, where the missing entries can be estimated using the RMC method described in the above iteration procedures.

**Fig. 1:**
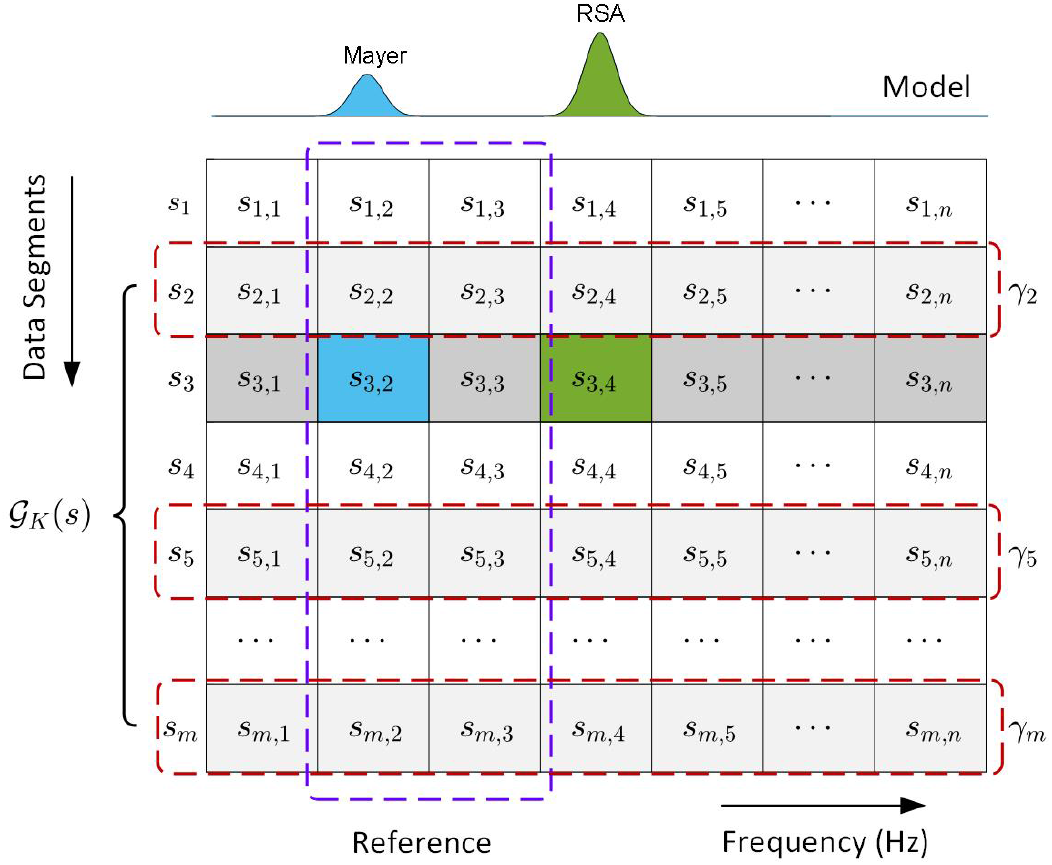
Illustration of matrix completion (MC) and the proposed refined matrix completion (RMC) using the model information for HRV spectrum estimation.

### D. Evaluation of Estimation Performance

The developed RMC method for HRV spectrum estimation is used to analyse a variety of datasets as discussed in the next section. To evaluate the model performance on spectrum estimation across different subjects and datasets, we use the normalised root mean square error (NRMSE) as the performance indicator [34]. For the estimation of missing entries in the **s**_*j*_ spectrum, the NRMSE index can be calculated as follows,

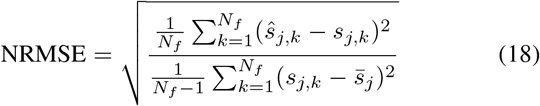

where, *s*_*j,k*_ *∈* **S** is the original spectrum, *j* = 1, 2, · · ·, *n*_*r*_ is the index of the spectrum, and *k* = 1, 2, · · ·, *N*_*f*_ indicates the data points of missing entries in the spectrum. ŝ_*j,k*_ is the estimated spectrum, and 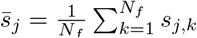 is the mean value of the spectrum. The calculation of this index normalises the root mean square error with the variance of the spectrum, which enables the evaluation of model performance across different ECG datasets using an indicator with the same scale.

## IV. Data Analysis

### A. Datasets

Generally, HRV data can be derived from ECG signals, and in the current study, we retrieved five widely used benchmark ECG datasets to evaluate the performance of our developed RMC model. These datasets are summarised as follows.

1. Combined measurement of ECG, Breathing and Seismocardiograms (*CEBSDB*) [35]: this dataset consists of ECG recordings sampled from 20 healthy volunteers. After excluding a low quality recording (‘m018’) due to the issue of electrode contact, the ECG signals collected from 19 subjects are used to for the study, which have a sampling duration of approximately 50 minutes.
2. MIT-BIH Supraventricular Arrhythmia Database (*SADB*) [36]: this dataset includes 78 ECG recordings sampled from subjects with supraventricular arrhythmias, and each signal has a recording time duration of 30 minutes.
3. St Petersburg INCART 12-lead Arrhythmia Database (*SPIADB*) [37]: this dataset consists of 75 ECG recordings with a duration of 30 minutes, which are collected from patients undergoing tests for coronary artery disease.

#### Algorithm 1

Estimation of HRV spectrum using the RMC method.

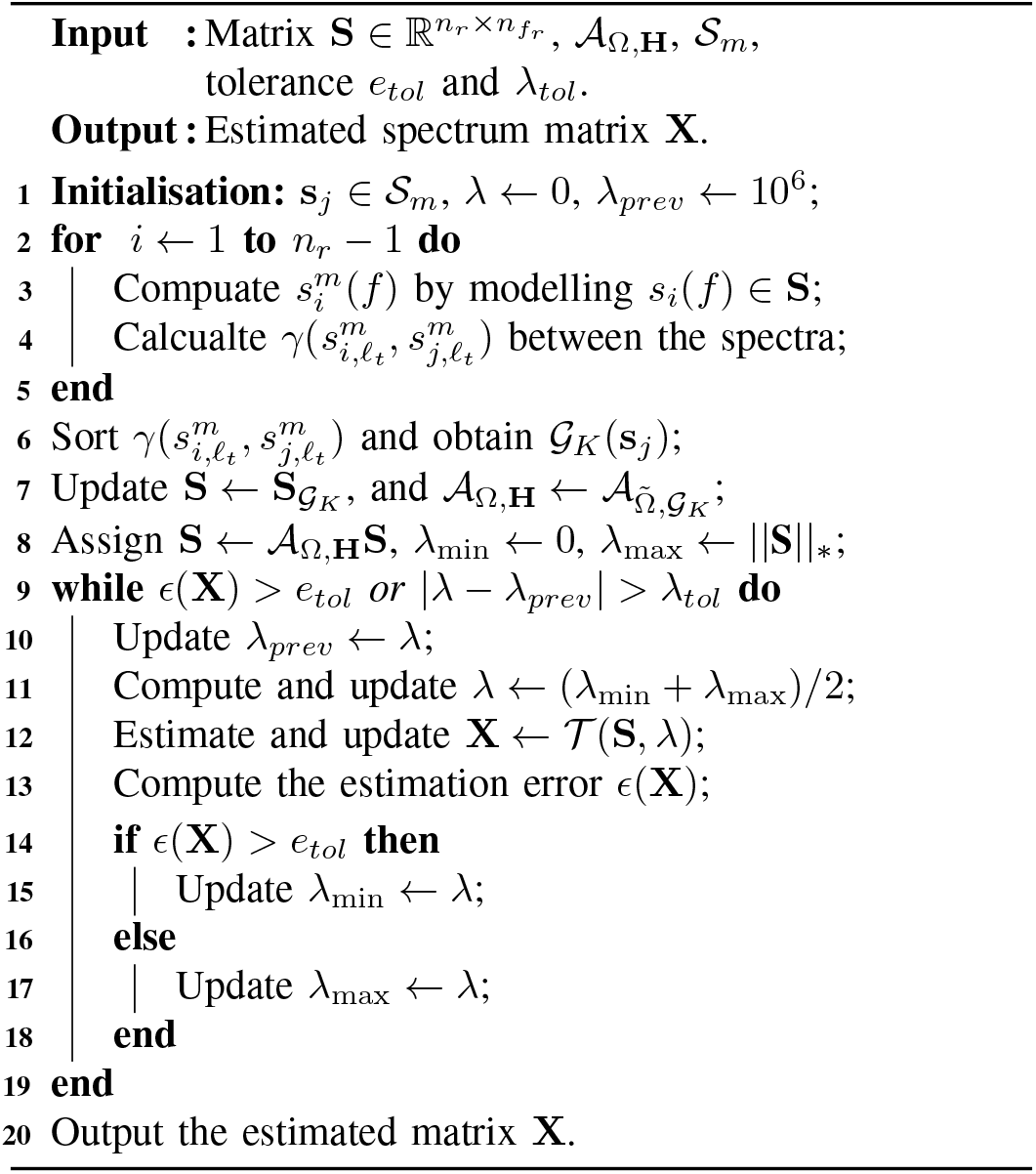
4. European ST-T Database (*ESTDB*) [38]: this database is widely used for the evaluation of ST and T-wave changes in the ECG morphologies, and includes 90 annotated excerpts of ambulatory ECG recordings with two-hour sampling durations.
5. Apnea-ECG Database (*APEDB*) [39]: this data consists of 70 ECG recordings with a set of apnea annotations, and each of the recordings has an approximate duration of 7 to 10 hours.

These datasets include ECG recordings with diverse characteristics, such as data sampled from healthy volunteers, patients with arrhythmias, coronary artery disease, ST and T-wave changes, and apnea. Therefore, the five ECG datasets will generate different types of HRV data, and in combination allow a comprehensive evaluation of our developed method for spectrum estimation. It is noted that the ECG signals in these datasets have varied lengths of sampling durations, and we use the first one hour of the signal only in instances of a long-time sampling duration.

### B. Signal Processing

To generate HRV data for this study, we first identify R-peaks from these ECG recordings by analysing the QRS complex. Figure 2(a) shows the identified R-peaks in the ECG recording using the QRS detector as developed in [40]. After identifying the R-peaks, a sequence of RR intervals can be calculated from timestamps of the consecutive peak values. Next, we implement outlier detection in the derived RR intervals using an open source benchmarked toolbox [41], where the outliers are defined as data points that are too close together, or large RR intervals caused by gaps, or artifact annotations of the signal. Figure 2(c) illustrates the detected outliers, which are processed by removal and interpolated to generate the preprocessed sequence.

**Fig. 2:**
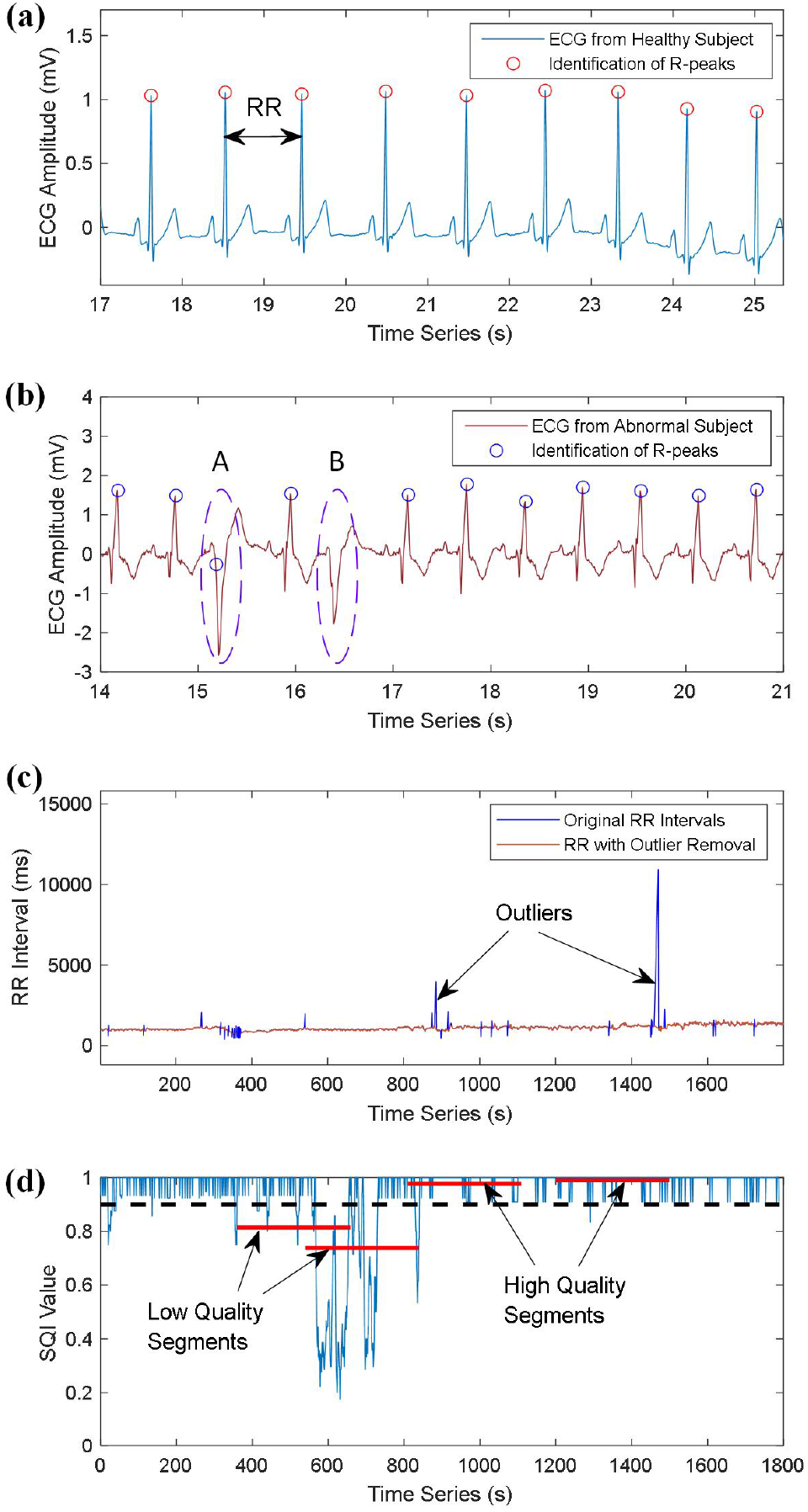
ECG signal processing and signal quality analysis of HRV data. (a) R-peaks identification for ECG recording sampled from healthy subject. (b) R-peaks identification for ECG recording sampled from abnormal subject. (c) Outlier detection in RR intervals. (d) Signal quality analysis for the HRV data.

We note that although a variety of techniques have been developed to detect the R-peaks [40], [42], it is still challenging to accurately identify the peak values for different types of ECG recordings. For example, as shown in Figure 2(a), the R-peaks are correctly identified for the ECG signal that is sampled from a healthy subject. However, as shown in Figure 2(b), the R peaks in waves *A* and *B* are wrongly identified for the ECG signal that is collected from an abnormal subject. The comparison of R-peak detection indicates that it may be not always reliable to use the technique to identify peak values for different types of ECGs. Therefore, a more robust approach needs to be developed to process the identified R-peaks.

To obtain high-quality HRV data, two widely used detectors are employed for the peak value identification in the study, i.e., the ‘jqrs’ detector [40], and the ‘wqrs’ detector [42]. Then, an index to show the signal quality (SQI) is computed for each of the identified R-peaks by comparing annotations from the two detectors [43]. Next, we calculate the average SQI value of R-peaks for HRV data, which is segmented with 5-minutes duration, and the calculated average value is used as a quality indicator for the data segment. For example, a high-quality segment can be defined when the SQI indicator is larger than 0.9; Otherwise, it is regarded as a low-quality segment. Figure 2(d) shows SQI values of the identified R-peaks in an ECG recording. As illustrated in Figure 2(d), the two data segments have SQI values of 0.813 and 0.737, and therefore are regarded as low-quality segments. By calculating the average SQI values, the low-quality and high-quality data segments can be efficiently identified, and the low-quality segments will not be used for further analysis.

### C. Calculation of the PSD Spectrum

The preprocessed HRV data is then resampled with a frequency of 4 Hz, and a 4^*th*^ order Butterworth filter is used to remove noises with the pass band of 0.03 Hz to 0.9 Hz [44]. The filtered data is then used to derive HRV spectrum in the frequency domain, and Welch’s algorithm with an overlap of 50% is used for the calculation [44], [45]. Next, the HRV spectrum matrix can be obtained by stacking all the calculated spectra. Figure 3(a) shows the three-dimensional plots of PSD spectra that are derived from different data segments. It can be seen from Figure 3(a) that the PSD spectra have similar patterns in the waveforms, i.e., LF and HF components, indicating the low-rank property of the HRV matrix derived from the PSD spectra.

**Fig. 3:**
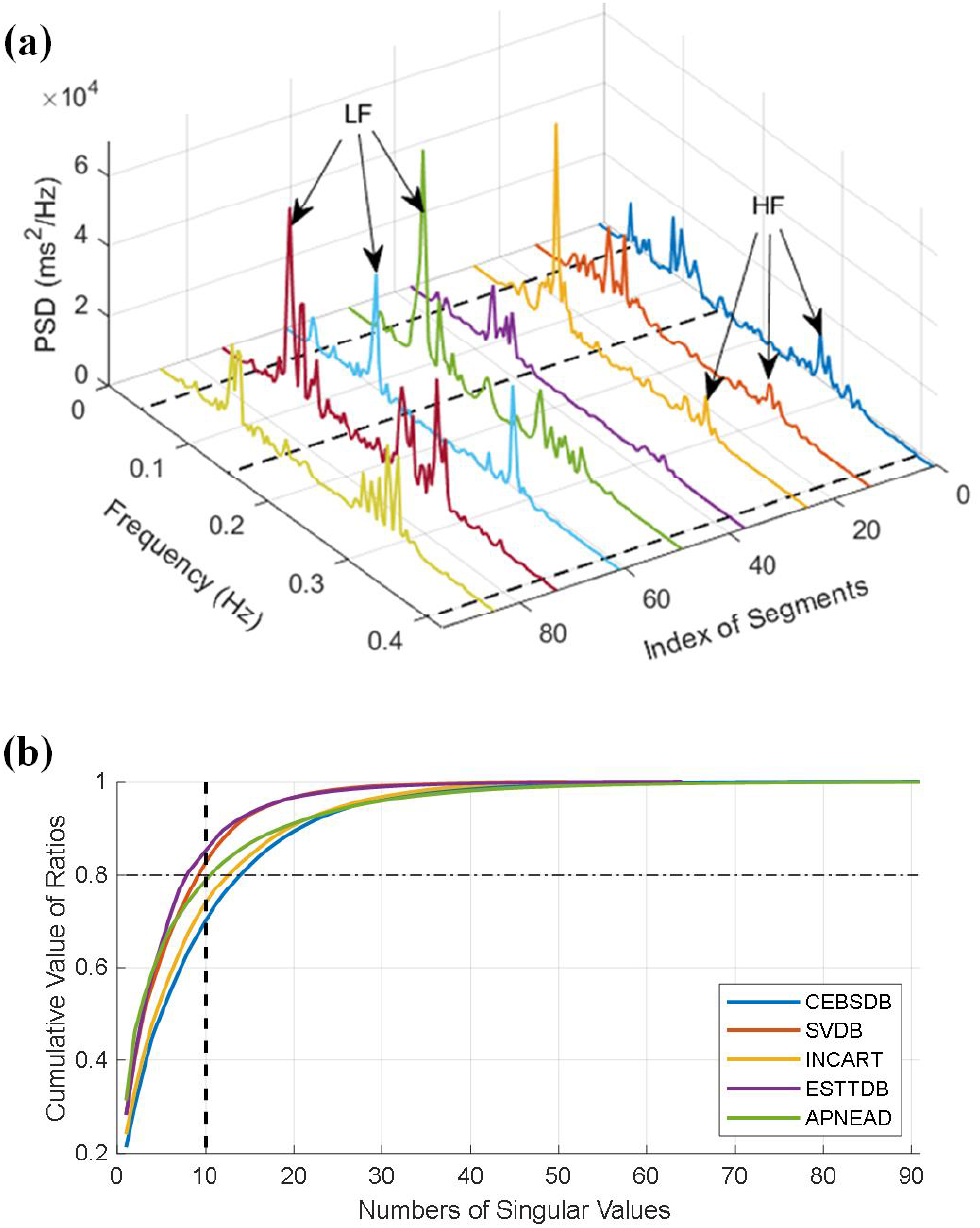
HRV spectrum and the cumulative ratios. (a) PSD spectra of different data segments. (b) Distributions of cumulative ratios between singular values of the nuclear norm.

In a further step, we calculate the singular values of the derived HRV matrix, and compute the ratio between singular values and the nuclear norm of the spectrum matrix, which is used as an indicator of the rank property of the matrix [34]. As shown in Figure 3(b), we demonstrate the calculated cumulative ratios for the five ECG datasets. It can be seen from the figure that the summation of the top ten singular values accounts for approximate 80% of the nuclear norm, indicating the low-rank properties of the spectrum matrices that are derived from these datasets.

## V. Results and Discussion

### A. Missing Data Estimation for PSD Spectrum

The derived HRV matrix is then used to evaluate the performance of our developed model for missing entry estimation, which is simulated by masking the spectra with different ratios. We show an example of masking 50% of the spectrum in Figure 4(a), which mostly represents the HF of the PSD spectrum. Then, the rest part of the spectrum, i.e., LF, is used to estimate the HF by the MC method. It can be seen from Figure 4(a) that the estimated HF spectrum matches well with the original PSD spectrum, which efficiently estimated the waveform for the HF of the spectrum but with large variations around the frequency of 0.25 Hz.

**Fig. 4:**
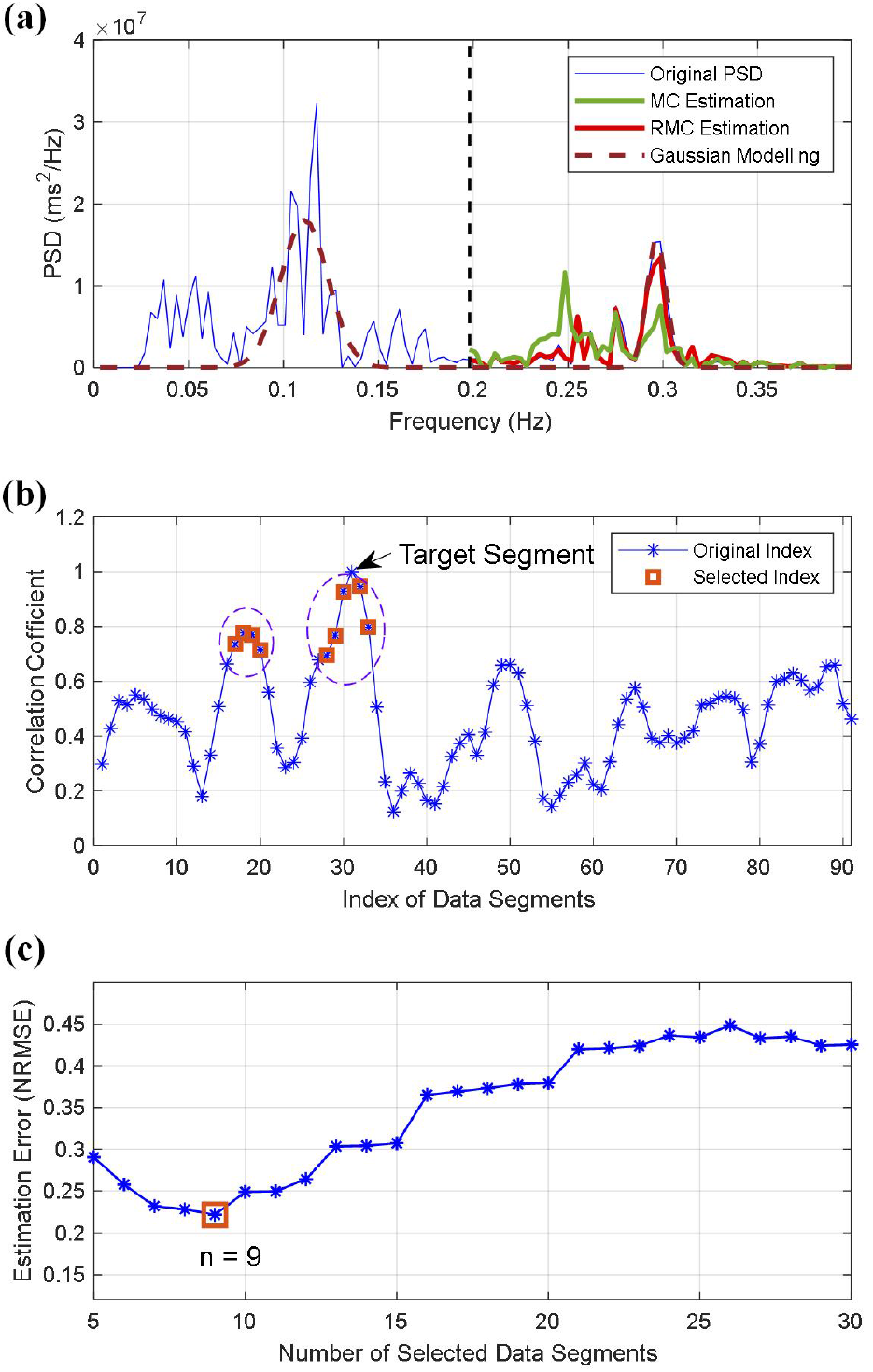
HRV spectrum estimation using matrix completion. (a) Spectrum estimation with different methods. (b) The correlation coefficients of different HRV segments. (c) Estimation errors of different combinations of data segments.

Next, we use the developed RMC method to estimate missing values in the spectrum. We first simulate the spectrum with the model as described in Eq. (13), and then use the modelled signal to identify relevant data segments in the HRV matrix. As illustrated in Figure 4(a), we show the signal modelling for the PSD spectrum of 31^*st*^ data segment of the matrix. It can be seen that the modelled signal efficiently represents the characteristics of the spectrum, such as the LF and HF. Then, we calculate correlation coefficients between different data segments using the modelled signals, and select the data with high coefficients to reconstruct the HRV matrix. Figure 4(b) shows the calculated coefficients for segments in the original data matrix, and Figure 4(c) illustrates estimation errors for different combinations of selected data segments.

It can be seen from Figure 4(c) that the RMC model obtains the least estimation error when using nine data segments, which has the value of 0.222 in terms of the NRMSE, and it is smaller than the estimation error of 0.830 using the traditional MC method. As shown in Figure 4(b), we highlight the selected data segments with square markers. It can be seen from Figure 4(b) that the selected important data elements include five neighbours of the target data segment, this is consistent with the rational approach that missing values can be generally imputed using nearest neighbours. We also note that the model identifies four segments as important data, which are not the neighbours of the target segment, indicating that some data with far distances in the sequence may also provide valuable information for the estimation of missing entries.

In a further step, we apply different masking ratios on the HRV spectrum, which are used to evaluate the model performance on estimating missing entries with different conditions. As shown in Figures 5 and 6, our developed RMC method efficiently estimates missing values for the PSD spectra with masking ratios of 30% (Figure 5(g)), and 70% (Figure 6(g)), which have estimation errors of 0.221 and 0.426 separately. The two experiments show the robustness of our developed RMC method for the estimation of missing entries in the HRV spectrum.

**Fig. 5:**
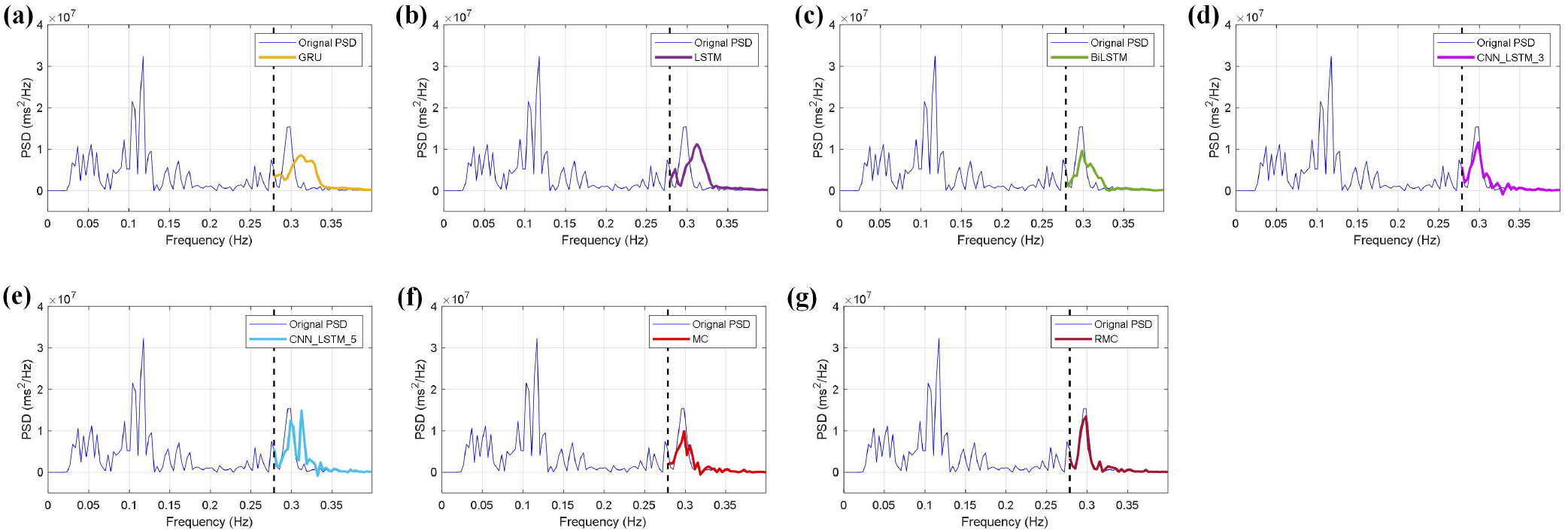
HRV spectrum estimation using different models for missing values with the 30% masking ratio. (a) The GRU model, (b) the LSTM model, (c) the BiLSTM model, (d) the CNN LSTM 3 model, (e) the CNN LSTM 5 model, (f) the MC method, and (g) the RMC method.

**Fig. 6:**
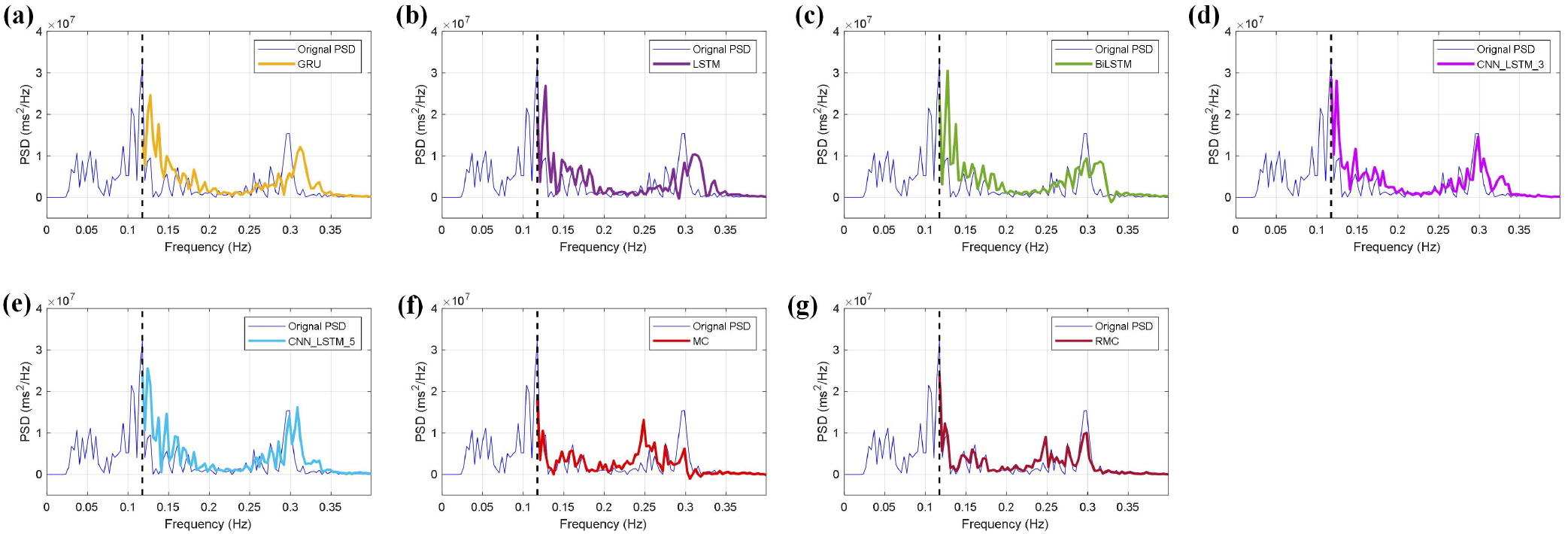
HRV spectrum estimation using different models for missing values with the 70% masking ratio. (a) The GRU model, (b) the LSTM model, (c) the BiLSTM model, (d) the CNN LSTM 3 model, (e) the CNN LSTM 5 model, (f) the MC method, and (g) the RMC method.

### B. Comparison Methods and Results

It is understood that machine learning techniques, in particular deep recurrent neural networks, have been demonstrated with excellent performance on prediction or regression analysis [46]. In this study, we employed different types of deep learning models to estimate missing values for comparison study. To be specific, we used the gated recurrent units (GRU), the long short-term memory (LSTM) model, and the bidirectional LSTM (BiLSTM). Moreover, we note that convolutional neural networks (CNN) are efficient in extracting features from data sequences [46]. Therefore, other than the GRU, LSTM, and BiLSTM, we used two additional hybrid models by combining the CNN and LSTM models for comparison study.

Hyperparameters of the five deep learning models are tuned with trial-and-error search, and the optimal parameters are obtained as follows; For the recurrent neural networks, the GRU, LSTM, and BiLSTM models have 120 hidden units; For the two types of hybrid models, the first one (CNN LSTM 3) uses three 1D-CNN layers with 32, 64, and 64 filters respectively, each filter has a size of 3, and each CNN layer is followed by a rectified linear unit (ReLU) function, and a bath normalisation layer, which is then flattened and connected with a LSTM layer for the regression analysis. The second type of hybrid models (CNN LSTM 5) consists of five CNN layers, which have 32, 32, 64, 64, and 64 filters respectively, which also uses a LSTM layer for the regression analysis. The deep learning models are trained with Adam optimizer with an initial learning rate of 0.005, the maximum epoch size is 100, and the batch size is set as 20. For the prediction analysis, we use data points from previous steps, i.e., 10 steps, to predict the current value in the data sequence.

The estimation results using these deep learning models are demonstrated in Figures 5 and 6, which correspond to the masking ratios of 30% and 70% respectively. As a comparison study, we also include the estimation results from the traditional MC method. It can be seen from Figures 5 and 6 that the deep learning models have good performance on the spectrum estimation; In particular, the BiLSTM and hybrid models efficiently estimate the peak values around 0.3 Hz for the two masking ratios. Compared with the deep learning models and the MC method, our developed RMC method has more efficient estimation performance, which obtains the least estimation errors of 0.221 and 0.426 for the 30% and 70% masking ratios respectively.

### C. Statistical Results

We implement the estimation of missing entries for all ECG recordings in the five benchmark datasets using our developed RMC method and the deep learning models. Tables I-III show the mean value and standard deviation of estimation errors for the five benchmark datasets with 30%, 50%, and 70% masking ratios. The minimum estimation error for each dataset is bold-faced. It can be seen from Tables I-III that compared with the five deep learning models and the traditional MC method, our developed RMC method obtains the least estimation error for all of the five ECG datasets, and the increased masks ratios occurred alongside an increase in the estimation errors, which have the values of 0.286 *±* 0.194, 0.312 *±* 0.203, and 0.379 *±* 0.228 for the CEBSDB with the three masking ratios.

**TABLE I:** Comparison of prediction errors for different estimation methods on benchmark datasets with 30% masking ratio.

| Datasets | GRU | LSTM | BiLSTM | CNN_LSTM_3 | CNN_LSTM_5 | MC | RMC |
| --- | --- | --- | --- | --- | --- | --- | --- |
| CEBSDB | $1.379 \pm 1.112$ | $1.346 \pm 0.970$ | $0.671 \pm 0.355$ | $0.672 \pm 0.459$ | $0.770 \pm 0.692$ | $0.393 \pm 0.291$ | **$0.286 \pm 0.194$** |
| SADB | $1.389 \pm 1.751$ | $1.318 \pm 1.280$ | $0.733 \pm 0.901$ | $0.746 \pm 0.902$ | $0.902 \pm 1.808$ | $0.332 \pm 0.253$ | **$0.304 \pm 0.199$** |
| SPIADB | $1.241 \pm 0.776$ | $1.211 \pm 0.609$ | $0.647 \pm 0.289$ | $0.655 \pm 0.375$ | $0.709 \pm 0.378$ | $0.361 \pm 0.539$ | **$0.305 \pm 0.327$** |
| ESTDB | $1.317 \pm 1.096$ | $1.297 \pm 1.043$ | $0.687 \pm 0.430$ | $0.691 \pm 0.456$ | $0.753 \pm 0.549$ | $0.494 \pm 0.464$ | **$0.329 \pm 0.251$** |
| APEADB | $1.283 \pm 0.740$ | $1.268 \pm 0.646$ | $0.644 \pm 0.263$ | $0.663 \pm 0.337$ | $0.727 \pm 0.392$ | $0.426 \pm 0.357$ | **$0.290 \pm 0.228$** |

**TABLE II:** Comparison of prediction errors for different estimation methods on benchmark datasets with 50% masking ratio.

| Datasets | GRU | LSTM | BiLSTM | CNN_LSTM_3 | CNN_LSTM_5 | MC | RMC |
| --- | --- | --- | --- | --- | --- | --- | --- |
| CEBSDB | $1.061 \pm 0.234$ | $1.091 \pm 0.275$ | $0.621 \pm 0.163$ | $0.619 \pm 0.164$ | $0.663 \pm 0.175$ | $0.432 \pm 0.192$ | **$0.312 \pm 0.203$** |
| SADB | $1.067 \pm 0.149$ | $1.093 \pm 0.190$ | $0.670 \pm 0.153$ | $0.673 \pm 0.129$ | $0.714 \pm 0.136$ | $0.386 \pm 0.254$ | **$0.338 \pm 0.226$** |
| SPIADB | $1.076 \pm 0.168$ | $1.097 \pm 0.201$ | $0.657 \pm 0.150$ | $0.662 \pm 0.115$ | $0.706 \pm 0.128$ | $0.388 \pm 0.529$ | **$0.310 \pm 0.290$** |
| ESTDB | $1.113 \pm 0.707$ | $1.138 \pm 0.692$ | $0.680 \pm 0.306$ | $0.704 \pm 0.427$ | $0.745 \pm 0.443$ | $0.590 \pm 0.446$ | **$0.336 \pm 0.276$** |
| APEADB | $1.059 \pm 0.292$ | $1.085 \pm 0.334$ | $0.642 \pm 0.216$ | $0.648 \pm 0.168$ | $0.687 \pm 0.175$ | $0.547 \pm 0.422$ | **$0.312 \pm 0.256$** |

**TABLE III:** Comparison of prediction errors for different estimation methods on benchmark datasets with 70% masking ratio.

| Datasets | GRU | LSTM | BiLSTM | CNN_LSTM_3 | CNN_LSTM_5 | MC | RMC |
| --- | --- | --- | --- | --- | --- | --- | --- |
| CEBSDB | $1.350 \pm 0.591$ | $1.446 \pm 0.676$ | $0.926 \pm 0.563$ | $0.906 \pm 0.451$ | $1.030 \pm 0.622$ | $0.633 \pm 0.202$ | **$0.379 \pm 0.228$** |
| SADB | $1.131 \pm 0.361$ | $1.247 \pm 0.422$ | $0.983 \pm 0.652$ | $0.819 \pm 0.238$ | $0.873 \pm 0.284$ | $0.539 \pm 0.318$ | **$0.397 \pm 0.262$** |
| SPIADB | $1.179 \pm 0.398$ | $1.325 \pm 0.544$ | $1.019 \pm 0.674$ | $0.828 \pm 0.230$ | $0.890 \pm 0.308$ | $0.502 \pm 0.406$ | **$0.338 \pm 0.247$** |
| ESTDB | $1.193 \pm 0.418$ | $1.335 \pm 0.543$ | $1.028 \pm 0.743$ | $0.849 \pm 0.304$ | $0.903 \pm 0.369$ | $0.728 \pm 0.356$ | **$0.337 \pm 0.252$** |
| APEADB | $1.144 \pm 0.368$ | $1.266 \pm 0.470$ | $0.958 \pm 0.689$ | $0.811 \pm 0.256$ | $0.864 \pm 0.315$ | $0.694 \pm 0.409$ | **$0.315 \pm 0.246$** |

Additionally, we provide violin plots as shown in Figure 7 to demonstrate the distributions of estimation errors for the seven models. It can be seen from Figure 7 that our developed RMC method obtains the least estimation errors with median values of 0.330, 0.337, 0.260, 0.259, and 0.235 for the five benchmark datasets respectively. In particular, we note that compared with the five deep learning models and the traditional MC method, the RMC method has much smaller variations of the estimation errors for all the ECG datasets, which indicate the stability and robustness of our developed RMC method for the spectrum estimation.

**Fig. 7:**
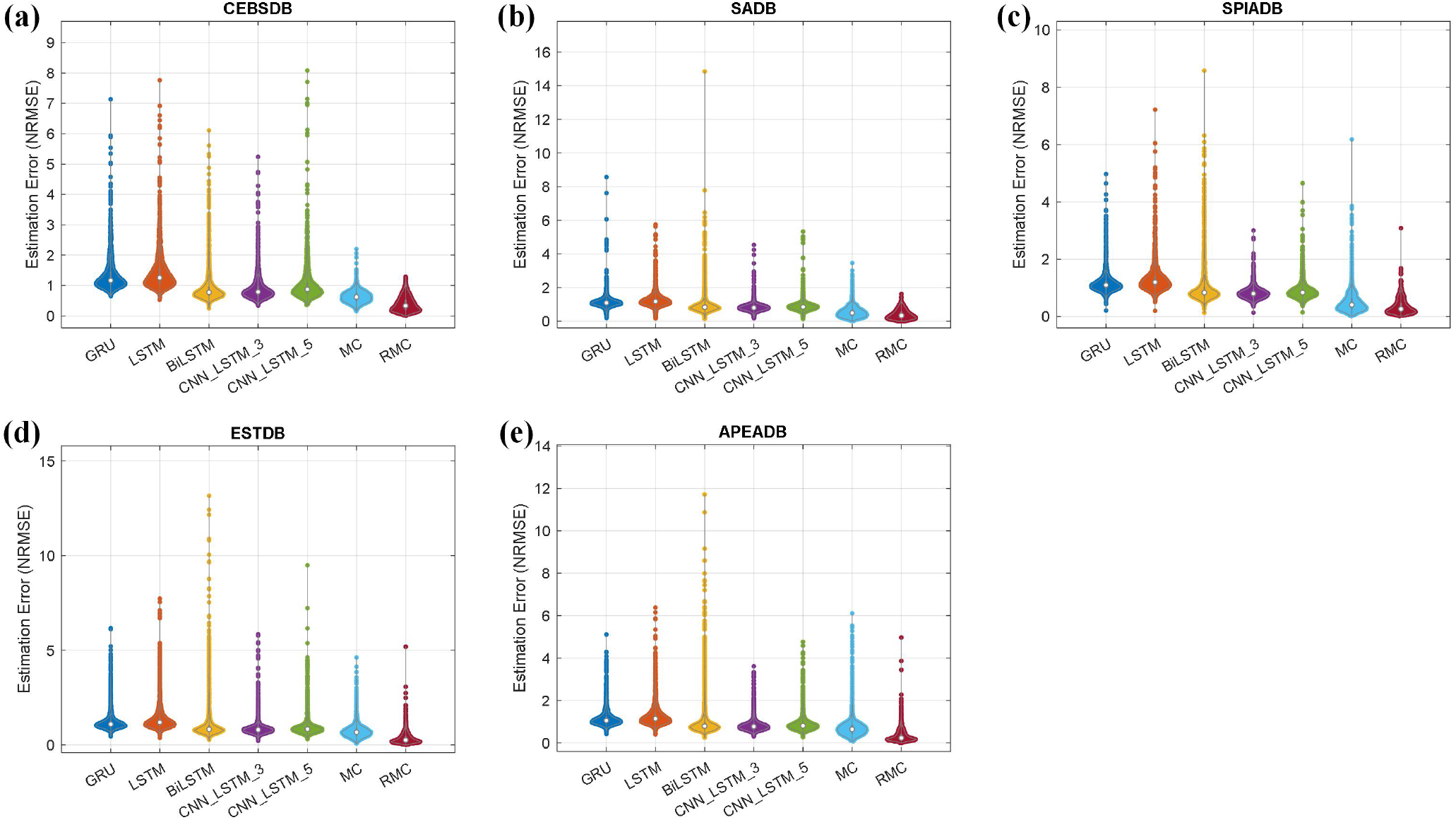
Distributions of estimation errors for missing values in five ECG benchmark datasets, (a) CEBSDB, (b) SADB, (c) SPIADB, (d) ESTDB, and (e) APEADB.

### D. Comparison on Computation Costs

We calculate the computation cost for missing entry estimation using the RMC method and other six methods. All of the estimation tasks are implemented with Matlab R2022a, Intel(R) Core(TM) i7-1165G7@2.8GHz, and 32GB RAM. We calculate the mean value and standard deviation of computation time for all HRV data segments in the benchmark dataset. Table IV presents the comparison of computation cost for all the estimation methods. We note that the five deep learning models have an increasing trend of computation cost with the increasing model complexity, which can be seen from Table IV that among these deep learning models, the GRU model has the least computation time with 1.117 *±* 0.460 s. Compared with the deep learning models and the MC method, our developed RMC method has the least computation time with 0.105 *±* 0.096 s. The results demonstrate the efficiency of our developed RMC method for the HRV spectrum estimation.

**TABLE IV:**
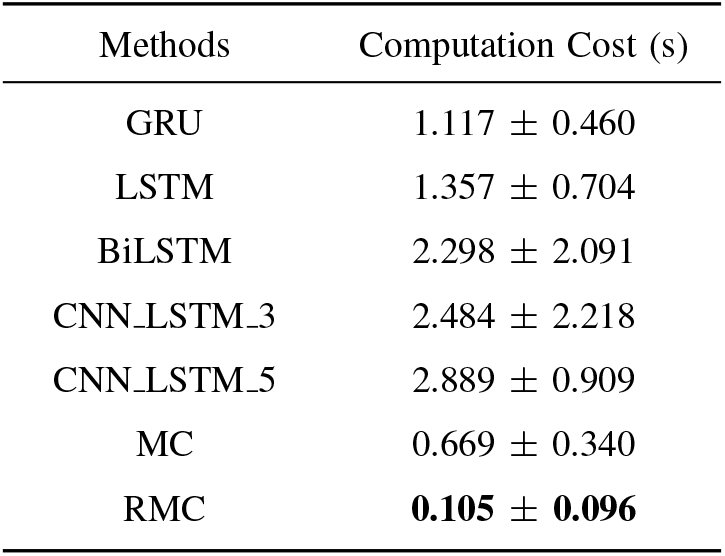
Comparison of computation time for different estimation methods.

### E. Discussion

HRV metrics - and in particular the spectrum parameters - are important indicators for physiological and pathological condition monitoring, such as the diagnosis of cardiovascular diseases [6], [7]. However, HRV metrics are sensitive to various measurement uncertainties in the datasets, such as motion artifacts and missing values. In this paper, we evaluate our developed method for missing entry estimation; this is crucial because HRV data can be easily affected by measurement noise and computational procedures, which would produce uncertainties in the spectrum analysis. For the sake of simplicity, we simulate the uncertainties as missing entries in the spectrum. With extensive experimental studies on five benchmark ECG datasets, we show that the PSD data that are derived from large amount of collected ECGs demonstrate similarities between the spectrum waveforms, which enables to use the MC technique for the spectrum estimation by exploring the low rank property of the matrix, we then show the advantages and robustness of our developed RMC method for the missing value estimation. Deep neural networks (DNN) have been demonstrated with excellent performance on analysing healthcare data for various applications [46]. However, DNN models typically require a large number of training data to ensure the model performance, which is a challenge when using small datasets. As demonstrated in the current study, we employed five widely used recurrent neural networks for comparison study, and the results demonstrated that our developed RMC method had superior performance than these deep learning models on all the benchmark datasets. The comparison results indicate that statistical machine learning techniques may have superior or comparable performance with deep learning models for small datasets. We also note that the MC method was developed with mathematical theory, our current study on the refined matrix completion was evaluated with extensive experimental studies, the next step of our research will focus on theoretical analysis of the developed RMC method.

## VI. Conclusions

This paper develops a new method to address the challenges of uncertainty in the estimation of HRV spectrum using the low-rank matrix completion method. By investigating the modelled spectrum of HRV data, a refined matrix completion (RMC) method was developed for the uncertainties of PSD spectrum. Five benchmark ECG datasets with three masking ratios of 30%, 50%, and 70% are used to evaluate the performance of our developed RMC method, and the results demonstrate the effectiveness and robustness of our developed method for the spectrum estimation. In addition, compared with five deep learning models and the traditional MC method, our developed RMC method obtains the least estimation error whilst having the least computation cost, indicating advantages and efficiency of our developed model for HRV spectrum estimation.

## Data Availability

All data produced are available online at https://physionet.org/about/database/

## Notes

This work was supported in part by the National Institute for Health Research (NIHR) Oxford Biomedical Research Centre (BRC), and in part by an InnoHK Project at the Hong Kong Centre for Cerebro-cardiovascular Health Engineering (COCHE). D. A. Clifton is an Investigator in the Pandemic Sciences Institute, University of Oxford, Oxford, UK. The views expressed are those of the authors and not necessarily those of the NHS, the NIHR, the Department of Health, InnoHK – ITC, or the University of Oxford. L. Lu is supported fully by InnoHK Project on Project 1.1 - Wearable Intelligent Sensing Engineering (WISE) at Hong Kong Centre for Cerebro-cardiovascular Health Engineering (COCHE).

### Competing Interest Statement

The authors have declared no competing interest.

### Funding Statement

This work was supported in part by the National Institute for Health Research (NIHR) Oxford Biomedical Research Centre (BRC), and in part by an InnoHK Project at the Hong Kong Centre for Cerebro cardiovascular Health Engineering (COCHE). D. A. Clifton is an Investigator in the Pandemic Sciences Institute, University of Oxford, Oxford, UK. The views expressed are those of the authors and not necessarily those of the NHS, the NIHR, the Department of Health, InnoHK ITC, or the University of Oxford. L. Lu is supported fully by InnoHK Project on Project 1.1 Wearable Intelligent Sensing Engineering (WISE) at Hong Kong Centre for Cerebro cardiovascular Health Engineering (COCHE).

### Author Declarations

The study used (or will use) ONLY openly available human data that were originally located at PhysioNet, https://physionet.org/about/database/

